# Selection into shift work is influenced by educational attainment and body mass index: A Mendelian randomization study

**DOI:** 10.1101/2020.03.10.20032698

**Authors:** Iyas Daghlas, Rebecca C. Richmond, Jacqueline M. Lane, Hassan S. Dashti, Hanna M. Ollila, Eva S. Schernhammer, George Davey Smith, Martin K. Rutter, Richa Saxena, Céline Vetter

## Abstract

**Background:** Shift work is associated with increased cardiometabolic disease risk, but whether this association is influenced by cardiometabolic risk factors driving selection into shift work is currently unclear. We addressed this question using Mendelian randomization (MR) in the UK Biobank.

**Methods:** We created genetic risk scores (GRS) associating with nine cardiometabolic risk factors (including education, body mass index [BMI], smoking, and alcohol consumption), and tested associations of each GRS with self-reported current frequency of shift work and night shift work amongst employed UKB participants of European ancestry (n=190,573). We used summary-level MR sensitivity analyses and multivariable MR to probe robustness of the identified effects, and tested whether effects were mediated through sleep timing preference.

**Results:** Genetically instrumented lower educational attainment and higher body mass index increased odds of reporting frequent shift work (odds ratio [OR] per 3.6 years [1-SD] decrease in educational attainment=2.40, 95% confidence interval [CI]=2.22-2.59, p=4.84 × 10^−20^; OR per 4.7kg/m^2^ [1-SD] increase in BMI=1.30, 95%CI=1.14-1.47, p=5.85 × 10^−05^). Results were unchanged in sensitivity analyses allowing for different assumptions regarding horizontal pleiotropy, and the effects of education and BMI were independent in multivariable MR. No causal effects were evident for the remaining factors, nor for any exposures on selection out of shift work. Sleep timing preference did not mediate any causal effects.

**Conclusions:** Educational attainment and BMI may influence selection into shift work, which may have implications for epidemiologic associations of shift work with cardiometabolic disease.

**Key messages:**

- Although it has been hypothesized that cardiometabolic risk factors and diseases may influence selection into shift work, little evidence for such an effect is currently available.
- Using Mendelian randomization, we assessed whether cardiometabolic risk factors and diseases influenced selection into or out of shift work in the UK Biobank.
- Our results were consistent with a causal effect of both higher BMI and lower educational attainment on selection into current shift work, with stronger effects seen for shift work that is more frequent and includes more night shifts.
- Using multivariable Mendelian randomization, we found that effects of higher BMI and lower education were independent. Sleep timing preference had a null effect on shift work selection and therefore did not mediate these effects.
- Selection through education and BMI may bias the relationship of shift work with cardiometabolic disease. Social mechanisms underlying these effects warrant further investigation.

## Introduction

Shift work is increasingly common and is a risk factor for cardiometabolic diseases^1^ including type 2 diabetes (T2D)^2^ and coronary artery disease (CAD)^3^. In these studies the baseline characteristics of shift workers often systematically differ from non-shift workers, including: higher rates of smoking^2,4,5^, lower rates of alcohol consumption^2,4^, lower educational attainment^6^, and greater body mass index (BMI)^6–8^. Such baseline differences may reflect either a selection effect^9^ of these factors on shift work, a causal effect of shift work on these factors, or confounding by shared influences. To date, few studies have examined evidence for a potential selection effect^10,11^.

Clarifying which factors influence participation in shift work is of public health interest. First, identifying factors that influence selection into shift work would offer basic insights into which populations are at highest risk for working in unfavorable conditions^12^. Second, shift work may mediate the effects of cardiometabolic risk factors, such as lower educational attainment^13^, on adverse health outcomes. Alternatively, factors influencing selection into shift work may confound observational relationships between shift work and adverse health outcomes. Estimating the magnitude of this selection effect could facilitate development of methodology to better account for this confounding and improve our understanding of the adverse effects of shift work on health.

Observational data have limitations for identifying selection factors, due to timing of exposure measurement, unmeasured confounding, and effects of shift work on the risk factor (i.e. reverse causation). One method to overcome these limitations is Mendelian randomization (MR)^14^. MR uses genetic variants as proxies for epidemiologic exposures to estimate causal effects. This approach is well-suited for identifying selection effects on shift work because genetic variants are precisely measured and less prone to bias through confounding or reverse causality^15,16^.

Previous MR analyses supported a causal effect of BMI on lower income and socioeconomic status^12,17^, but MR has not been applied to characterize the determinants of selection into shift work. We used MR to assess the causal effect of cardiometabolic risk factors and diseases on shiftwork selection in the UK Biobank^18^.

## Methods

### Population

The UK Biobank is a population-based cohort study that enrolled over 500,000 volunteers aged 40-69 from 2006-2010; participation rate: 5.5%^18^. Data collection included questionnaire and nurse interview information, anthropometric and physiological measurements, and genomic data as previously described^18,19^. All participants provided written informed consent, and data used in this study were de-identified.

### Exposures

The exposures were single genetic variants or weighted genetic risk scores (GRS) derived from genome-wide association studies (GWAS) for cardiometabolic risk factors and diseases: alcohol consumption, smoking heaviness, body mass index (BMI), waist-to-hip ratio adjusted for BMI, type 2 diabetes, coronary artery disease, and educational attainment (Table 1)^20,21,22,23,24,13,25,26,27^. We focused on risk factors with publicly available GWAS summary statistics, and with putative causal effects on cardiometabolic outcomes, as supported by prior MR studies^31,20,28–31^. A single nucleotide polymorphism (SNP) in the alcohol dehydrogenase 1B (*ADH1B*) gene robustly associated with reduced alcohol consumption^20^ was used to proxy alcohol consumption. Consistent with previous analyses^20^, we coded this missense variant (rs1229984) under a dominant model (cases combining homozygotes and heterozygotes), with the effect allele oriented to reduced alcohol consumption. A single missense variant (rs16969968) in the Cholinergic Receptor Nicotinic Alpha 5 Subunit (*CHRNA5*) gene, associated at genome-wide significance with one additional cigarette smoked per day^21^, was used as the genetic instrument for smoking heaviness (limiting the sample to ever smokers^32^). For all other exposures, we used SNP associations from GWAS meta-analyses that did not include UKB data to generate multi-SNP GRS^22,23,25,27^. Figure 1 shows a causal diagram by which these exposures may influence shiftwork selection.

**Table 1.**
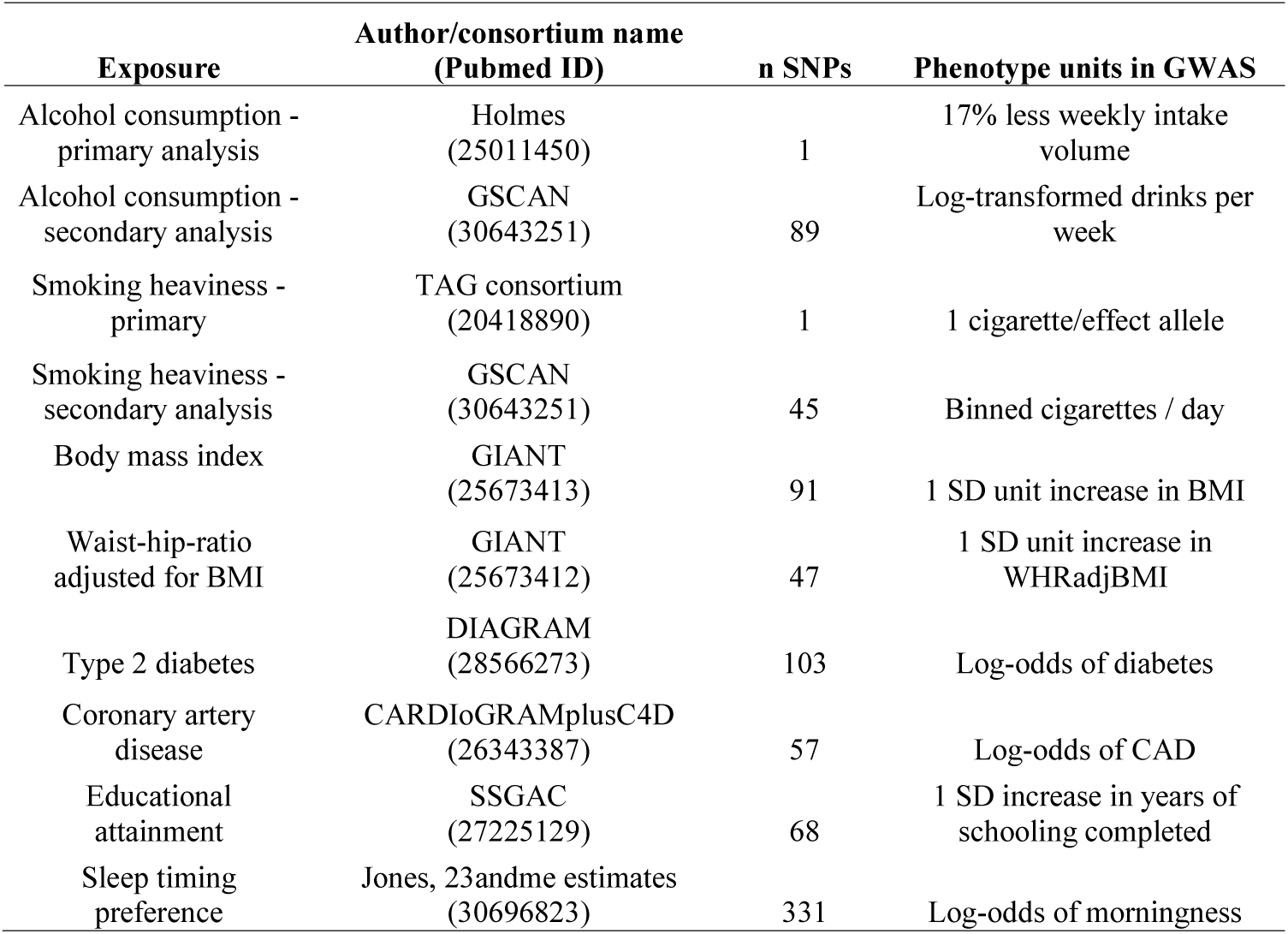
Sources of SNP-exposure associations for genetic risk scores.

| **Exposure** | **Author/consortium name**<br>**(Pubmed ID)** | **n SNPs** | **Phenotype units in GWAS** |
| --- | --- | --- | --- |
| Alcohol consumption -<br>primary analysis | Holmes<br>(25011450) | 1 | 17% less weekly intake<br>volume |
| Alcohol consumption -<br>secondary analysis | GSCAN<br>(30643251) | 89 | Log-transformed drinks per<br>week |
| Smoking heaviness -<br>primary | TAG consortium<br>(20418890) | 1 | 1 cigarette/effect allele |
| Smoking heaviness -<br>secondary analysis | GSCAN<br>(30643251) | 45 | Binned cigarettes / day |
| Body mass index | GIANT<br>(25673413) | 91 | 1 SD unit increase in BMI |
| Waist-hip-ratio<br>adjusted for BMI | GIANT<br>(25673412) | 47 | 1 SD unit increase in<br>WHRadjBMI |
| Type 2 diabetes | DIAGRAM<br>(28566273) | 103 | Log-odds of diabetes |
| Coronary artery<br>disease | CARDIoGRAMplusC4D<br>(26343387) | 57 | Log-odds of CAD |
| Educational<br>attainment | SSGAC<br>(27225129) | 68 | 1 SD increase in years of<br>schooling completed |
| Sleep timing<br>preference | Jones, 23andme estimates<br>(30696823) | 331 | Log-odds of morningness |

**Figure 1.**
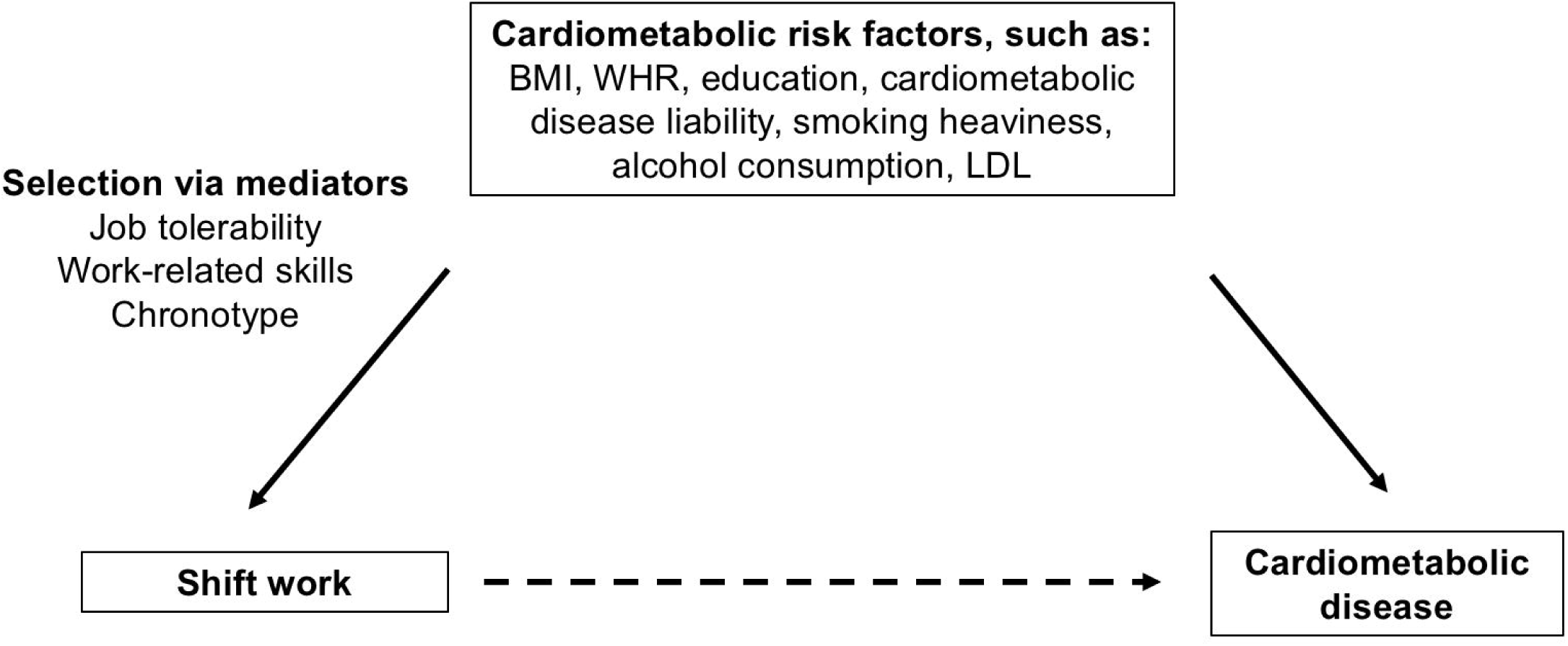
Causal diagram by which shift work may influence cardiometabolic risk. Shift may directly cardiometabolic risk, however selection into shift work through cardiometabolic risk factors or disease may also induce or modify relationships.

We utilized risk profiling in PLINK^33^ v1.9 to construct beta-weighted GRS using individual-level data in the UKB for participants of White British ancestry; weights were obtained from the respective GWAS. We regressed each GRS on the respective exposure to determine the strength of each genetic instrument, using an F-statistic >10 to indicate minimal influence of weak instrument bias^34^.

### Outcomes

We derived the study outcomes using current shift work characteristics at baseline from currently employed UKB participants. Participants were asked to indicate shift work participation by answering the following question: *‘Does your work involve shift work?’ defined as ‘a work schedule that falls outside of the normal daytime working hours of 9am-5pm. This may involve working afternoons, evenings or nights or rotating through these kinds of shifts*.*’* Response options included: *‘never/rarely,’ ‘sometimes,’ ‘usually,’ ‘always,’ ‘prefer not to answer,’ and ‘do not know*.*’ ‘Never/rarely’* served as the reference group, *‘sometimes’* was an intermediate group, and *‘usually’* and *‘always’* were collapsed into *‘frequent shift work;’* totaling three groups in this analysis. *‘Prefer not to answer’* and *‘do not know’* were coded as missing.

Participants indicating that they currently worked shifts were further queried, *‘Does your work involve night shifts,’ defined as ‘a work schedule that involves working through the normal sleeping hours, for instance working through the hours from 12am to 6am*.*’* Response options were identical to those for the primary shift work question and coded analogously. In the night shift work analyses we compared employed individuals not participating in shift work (reference group) with individuals working *‘shift work, but no night shift work,’ ‘some night shift work,’* and *‘frequent night shift work;’* resulting in four groups in this analysis.

All UKB participants with email addresses (n∼330,000) were invited to complete an online follow-up questionnaire on lifetime employment information. This questionnaire queried information on each job worked over the participant lifetime (n=118,699). For each job, participants were asked *‘Did you ever work shifts (day and/or night shifts) for this job?’*. To determine whether associations generalized beyond current shiftwork participation, we used this variable to derive an outcome of lifetime history of shiftwork, regardless of current employment status (n=83,133; 22,987 ever shift workers / 60,146 never shift workers). To identify exposures influencing selection out of shift work, we tested the outcome of history of quitting shift work. Cases were defined as participants reporting working a shiftwork job and subsequently working a non-shift work job (n=22,987; 14,584 cases of participants who ever quit shift work / 8,403 controls who worked shifts and never quit) as shown in Figure 2.

**Figure 2.**
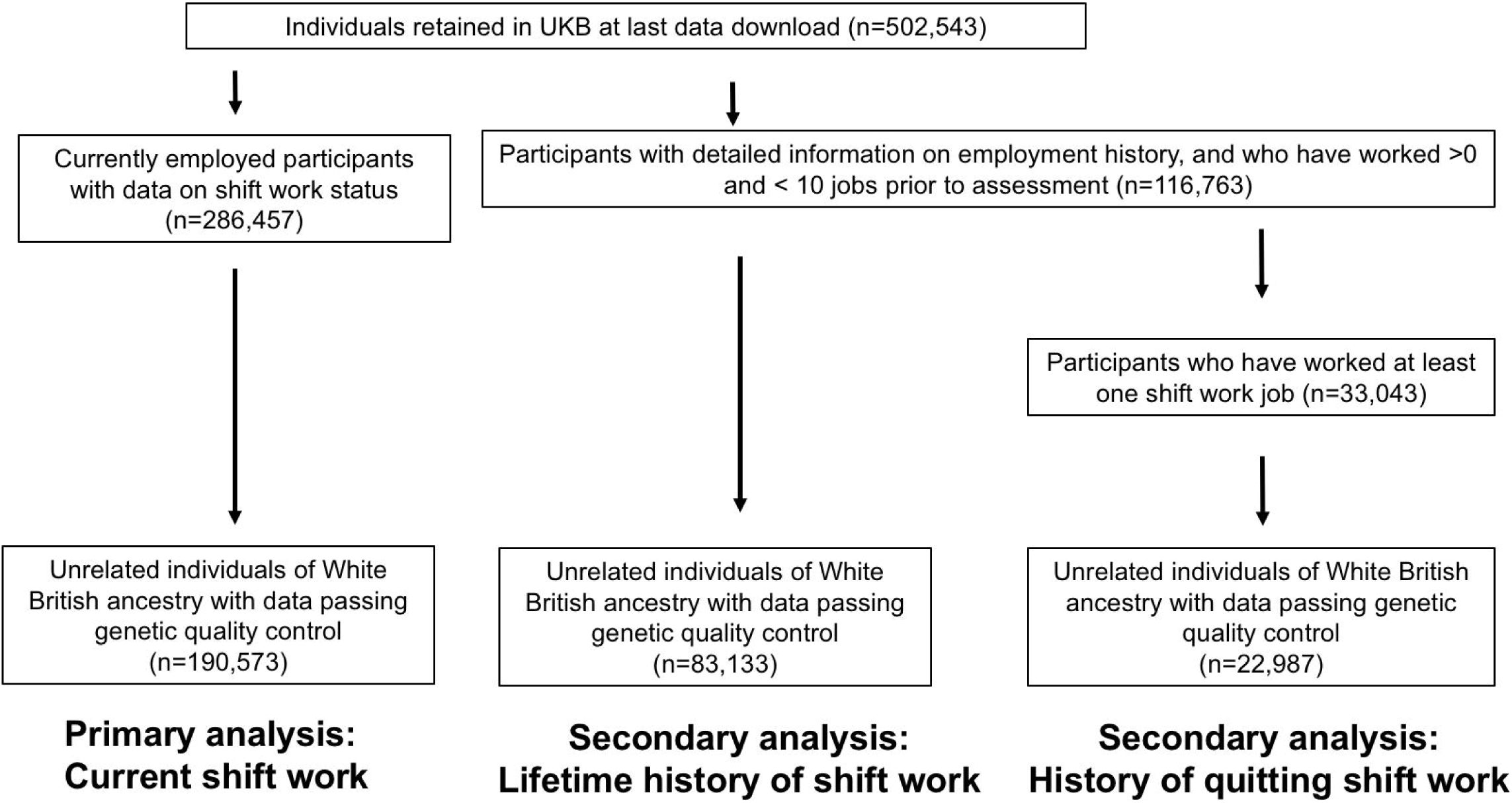
Analysis workflow.

### Individual-level analyses

We first tested the association of each GRS^35^ with categories of the shiftwork outcomes using multinomial or binomial logistic regression adjusted for age, sex, and the top ten principal components (PCs) of ancestry. We transformed the effects of binary exposures (GRS for CAD and T2DM) to reflect a doubling in the odds of the exposure on the odds of the outcome^36^ (i.e. the effect of lifelong increased risk for disease on shift work selection).

We then probed the robustness of results through a series of secondary and sensitivity analyses. In order to include individuals who previously worked a job with shifts, but would no longer be included in the “current shift work” analysis, we also tested associations with lifetime history of shiftwork participation. To identify exposures influencing selection out of shift work, we tested associations with history of leaving shiftwork. To assess the impact of population stratification^37^, we controlled for i) 40 PCs of ancestry, and ii) the 22 UK Biobank assessment centers. Prior work suggested sex differences in the causal effects of BMI on socioeconomic outcomes^17^, so we tested for GRS interactions with sex using a model with an interaction term. We used UKB job codes to stratify jobs as ‘skilled’ and ‘unskilled’, using the classification laid out in Howe et al^12^, and tested for interactions of job skill with the GRS using a model with an interaction term.

Finally, we expanded the alcohol consumption and smoking heaviness instruments to include SNPs identified in a recent large-scale meta-analysis^38^. We used GRS weights from summary statistics excluding UKB^38^. These instruments explain more variance in the exposures relative to the single SNP instruments, but may be at greater risk for pleiotropy due to the inclusion of multiple variants with uncharacterized function^39^.

### Summary-level MR analyses and sensitivity analyses

Genetic associations may be explained by horizontal pleiotropy, which is when variants influence the outcome through paths independent of the exposure of interest. We therefore conducted summary-level MR analyses to further assess whether the genetic associations represented casual relationships. We first estimated causal effects using inverse-variance weighted (IVW) random-effects regression, which is robust to balanced pleiotropy^40^. As a global test for pleiotropy, we calculated Cochran’s Q for heterogeneity. We then used the following pleiotropy-robust sensitivity analyses: MR Egger^41^, weighted median^42^, and MR-PRESSO^43^ regressions (see Supplementary methods 1). We used the estimate of the MR Egger model intercept to further assess for balanced horizontal pleiotropy. Due to low power^44^, this method is generally more appropriate for ruling pleiotropy in rather than ruling it out.

These analyses were implemented using the TwoSampleMR R package v0.4.22^45^. For effects identified in univariable MR, we undertook summary-level multivariable MR (MVMR)^45^ simultaneously controlling for each exposure using the TwoSampleMR software^45^. We confirmed instrument strength using the Q_strength_ adapted for MVMR^46^. The Q_strength_ divided by the number of variants is analogous to the F statistic, with values >10 indicating minimal weak instrument bias. Prior MR analyses identified a nominal effect of greater BMI on earlier sleep timing preference so we tested for a causal effect of early sleep timing preference on selection into shift work using GWAS estimates that did not overlap with UKB^47^. We hypothesized that earlier sleep timing preference would be negatively associated with current shift work, reflecting a negative selection effect. We planned to test MVMR of the exposures and sleep timing preference only if the first-stage association of sleep timing preference with shift work demonstrated an effect.

### Statistical software

Analyses were conducted using R version 3.5.0, including the TwoSampleMR^45^ and MVMR^46^ packages.

## Results

### Individual level analyses

The median age of employed participants in the UKB was 53 years [IQR 47-59], and 49% were male. Shift workers were more likely to be male and report lower indicators of socioeconomic and health status (Table 2). Each GRS was strongly associated with the exposures (Supplementary Table 1).

**Table 2.** Sample characteristics by current shift work status (n=190,569). Summary statistics are shown as mean (standard deviation) or count (percentage).

| **Exposure or covariate** | **Employed, not**<br>**shift worker**<br>**(n=159,900)** | **Some shift work**<br>**(n=13,411)** | **Frequent shift work**<br>**(n=17,258)** |
| --- | --- | --- | --- |
| Age (years) | 53.13 (7.08) | 52.34 (7.00) | 51.94 (6.87) |
| Male sex (%) | 75794 (47.4) | 7477 (55.8) | 9692 (56.2) |
| Household income < 18,000 (%) | 12898 (8.9) | 1496 (12.4) | 2295 (14.7) |
| Household income > 100,000 (%) | 12056 (8.3) | 641 (5.3) | 268 (1.7) |
| University education | 63824 (39.9) | 3749 (28.0) | 2433 (14.1) |
| Years of education | 16.01 (4.70) | 15.40 (4.82) | 14.55 (4.78) |
| Townsend deprivation index | -1.72 (2.79) | -1.10 (3.06) | -0.83 (3.12) |
| Lives with spouse or partner (%) | 121599 (83.3) | 9486 (79.6) | 11844 (77.9) |
| Current smoker (%) | 15002 (9.4) | 1928 (14.4) | 2745 (16.0) |
| Drinks per week (standardized<br>alcohol units) | 15.78 (16.02) | 16.49 (17.32) | 14.61 (16.61) |
| BMI (kg/m<sup>2</sup>) | 27.08 (4.62) | 27.94 (4.89) | 28.14 (4.91) |
| Waist-hip-ratio | 0.86 (0.09) | 0.88 (0.09) | 0.89 (0.09) |
| LDL (mmol/L) | 3.67 (0.82) | 3.70 (0.85) | 3.68 (0.83) |
| Coronary artery disease (%) | 2130 (1.3) | 227 (1.7) | 289 (1.7) |
| Type 2 diabetes (%) | 3757 (2.3) | 379 (2.8) | 529 (3.1) |
| Early sleep timing preference<br>(%)<sup>1</sup> | 36,572 (25.5) | 3,230 (26.9) | 3,823 (25.1) |
| Excellent self-rated health (%)<sup>2</sup> | 31588 (19.8) | 2019 (15.1) | 2356 (13.7) |
| Poor self-rated health (%)<sup>2</sup> | 3084 (1.9) | 336 (2.5) | 544 (3.2) |

<sup>1</sup> Scale for sleep timing preference includes: “Definitely a ‘morning’ person,” “More a ‘morning’ than ‘evening’ person,” “More an ‘evening’ than a ‘morning’ person,” and “Definitely an ‘evening’ person.”
<sup>2</sup> Scale for self-rated health includes: poor, fair, good, and excellent.

Genetically predicted educational attainment increased odds of working some shift work (odds ratio per 1-standard deviation (SD) / 3.6y (OR) 1.32, 95% confidence interval (CI) 1.10-1.59, p=2.64×10^−03^), frequent shift work (OR=2.22, 95%CI=1.89-2.63, p<2 × 10^−16^), and frequent night shift work (OR 2.94, 95%CI=2.27-3.85, p< 2×10^−16^; Figure 4). (Figures 3-4). Genetically predicted BMI increased odds of working ‘some’ shift work (OR 1.21, 95% CI 1.08-1.35, p=9.20 × 10^−04^), ‘frequent’ shift work (OR 1.25, 95% CI 1.14-1.38, p=7.40 × 10^−06^), and frequent night shift work (OR 1.44, 95% CI 1.23-1.68, p=4.00 × 10^−06^; Figures 3-4).

**Figure 3.**
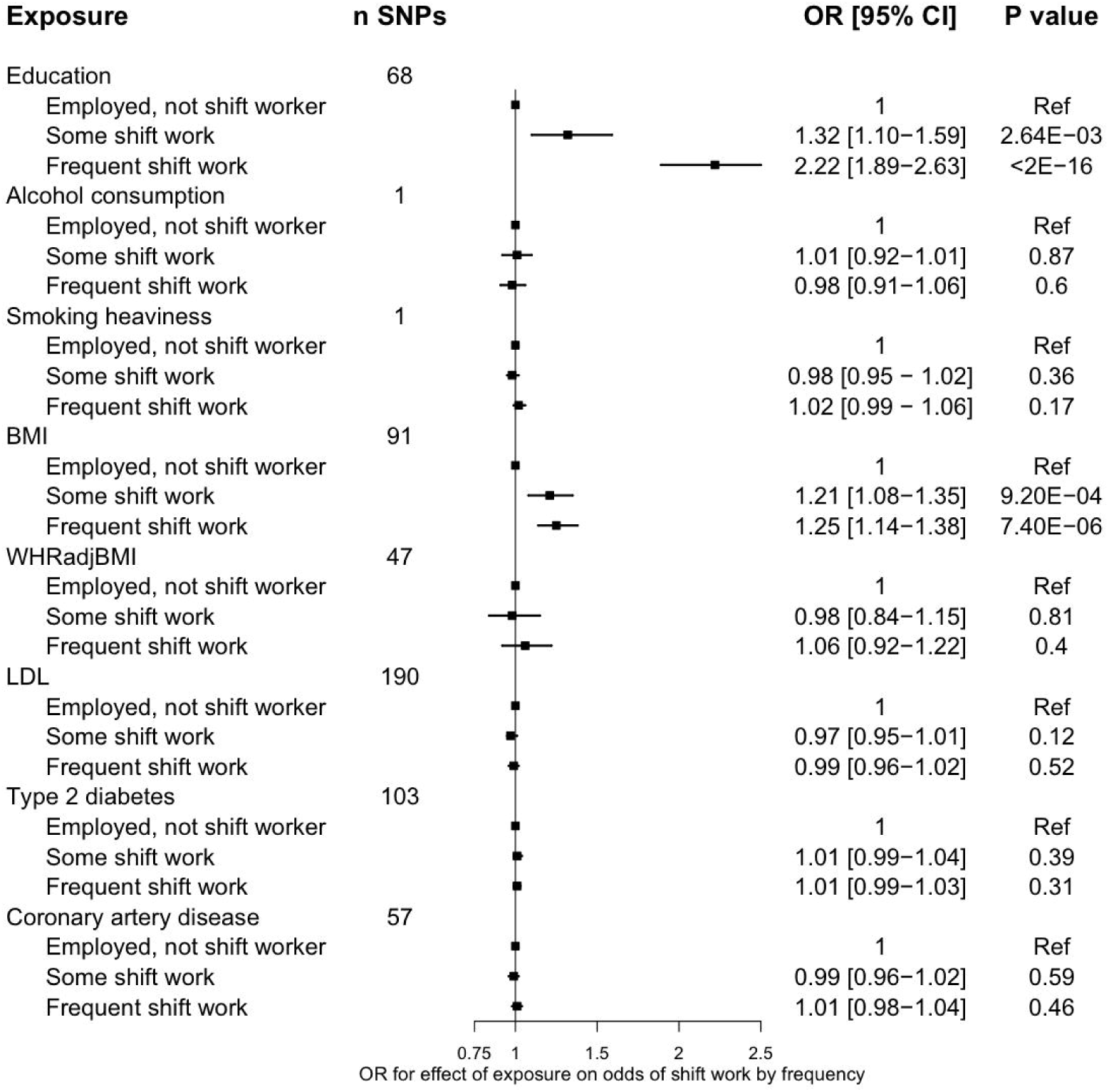
Associations of genetic instruments for cardiometabolic traits with current overall shift work^1^. The effect sizes correspond to a unit change in the GRS, or to an additional effect allele for the single-SNP instruments. ^1^n_total_=190,573; n_controls_=159,903, n_some shift work_=13,411, n_freqnent shift work_=17,259 BMI: body mass index; CAD: coronary artery disease; CI: confidence interval; GRS: genetic risk score; SNP: single nucleotide polymorphism; DM: diabetes mellitus; OR: odds ratio; WHR: waist-to-hip ratio adjusted for BMI

**Figure 4.**
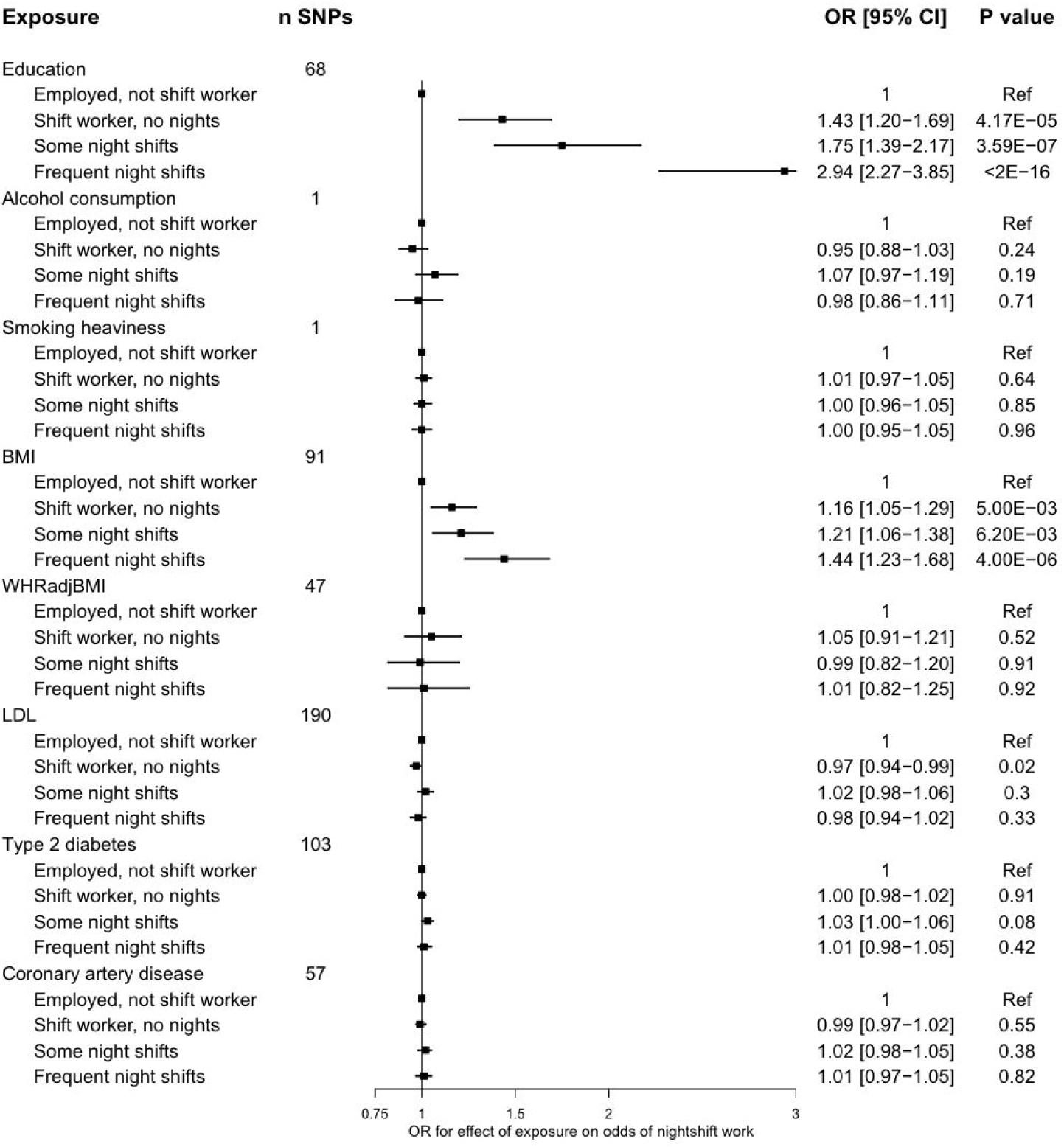
Associations of genetic instruments for cardiometabolic risk factors and disease with current night-shift work^1^. The effect sizes correspond to a unit change in the GRS, or to an additional effect allele for the single-SNP instruments. ^1^ntotal=190,573, n_controls_=159,903, n_non-night shift_=15,288, n_some night shift_=8,726; n_frequent night sfhit_=6,629) BMI: body mass index; CAD: coronary artery disease; CI: confidence interval; GRS: genetic risk score; SNP: single nucleotide polymorphism; DM: diabetes mellitus; WHR: waist-to-hip ratio adjusted for BMI

Both educational attainment (OR 1.40, 95% CI 1.20-1.64, p=2.68 × 10^−05^) and BMI (OR 1.17, 95% CI 1.06-1.28, p=1.68 × 10^−03^) increased the odds of working at least one shiftwork job across the life course (Supplementary Table 2). No other exposures associated with shift work participation (Figures 3-4). No exposures were associated with history of leaving shift work (Supplementary Table 3).

### Secondary and sensitivity analyses in individual level analyses

The BMI and education GRS associations with shift work did not interact with sex or job skill classification. Additional adjustment for population stratification through controlling for 40 principal components and UKB assessment center did not influence the BMI or education effects (Supplementary Table 4). Results were similar when using an expanded GRS for alcohol consumption (OR=1.07, 95%CI=0.88-1.32, p=0.50) and smoking heaviness (OR=1.03, 95%CI=1.00-1.05, p=0.02) on the outcome of frequent shift work. There was no effect of genetic propensity to early sleep timing preference on frequent shift work (OR 0.99, 95% CI 0.96-1.04, p=0.79) or frequent night shift work (OR 0.96, 95% CI 0.90-1.02, p=0.16).

### Summary-level MR analyses

We conducted summary-level MR analyses to estimate instrumental causal effects robust to horizontal pleiotropy. IVW random-effects models showed consistent effects of higher BMI (OR 1.30, 95% CI 1.14-1.47, p=5.58×10^−05^) and lower educational attainment (OR 2.40, 95% CI 2.22-2.59, p=4.84×10^−20^) on shift work (Table 3). Four lines of evidence suggest that the observed effects were not explained by horizontal pleiotropy. First, there was minimal evidence of heterogeneity (Table 3). Second, similar MR effects were observed across the Egger and weighted median analyses (Table 3). Third, there was no evidence of unbalanced pleiotropy as estimated by the Egger intercepts for the effects of BMI (intercept=-0.004, p=0.37) and of education (intercept=0.015, p=0.07) on shift work, although this test is generally underpowered. Fourth, MR-PRESSO did not detect pleiotropic outlier SNPs.

**Table 3.** Summary-level MR analyses relating educational attainment and BMI to shift work status^1^.

| Exposure | MR technique |  |  |  |  |  |  |  |
| --- | --- | --- | --- | --- | --- | --- | --- | --- |
|  | Inverse-variance weighted |  | MR Egger |  | Weighted median |  | Heterogeneity statistic |  |
|  | OR<br>[95% CI] | P value | OR<br>[95% CI] | P value | OR<br>[95% CI] | P value | Q | P value |
| Education | 2.40<br>[2.22, 2.59] | 4.84E-20 | 5.26<br>[2.22, 12.5] | 3.30E-04 | 2.22<br>[1.72, 2.94] | 1.01E-09 | 76 | 0.13 |
| BMI | 1.30<br>[1.14, 1.47] | 5.58E-05 | 1.51<br>[1.05, 2.16] | 0.027 | 1.34<br>[1.13, 1.59] | 8.68E-04 | 126 | 0.01 |

<sup>1</sup>n= 17,259 cases of 'frequent shift work' / 159,903 controls
adj.: adjusted for; OR: odds ratio; CI: confidence interval; MVMR: multivariable MR

We next turned to multivariable MR to assess the independence of exposure effects on shift work. The genetic instruments for educational attainment (Q_strength_=2,407, F_adjusted_=15.4) and BMI (Q_strength_=4,393, F_adjusted_=28) were conditionally strong in multivariable MR. The effects of BMI (OR=1.18, 95%CI=1.08-1.31, p=1.94×10^−03^) and of education (OR=2.56, 95%CI=2.12-3.13, p=1.48×10^−23^) on self-reported ‘frequent’ shift work were independent in MVMR.

## Discussion

These analyses revealed a causal and independent effect of lower educational attainment and higher BMI on current and lifetime selection into shift work, particularly night shift work. These effects were not mediated by sleep timing preference. There was minimal evidence for selection through other tested exposures.

Our findings provide evidence for a causal effect of lower educational attainment and higher BMI, but not of other cardiometabolic risk factors, on selection into shift work within this population. The effect sizes were larger when shift work involved night shifts, and with increasing frequency of shift work, supporting a dose-dependent effect. Selection into shift work (including alcohol consumption, BMI, smoking, lipids, and glucose) by cardiometabolic risk factors has been evaluated in previous observational studies (N∼2,800) ^10,11^. These studies identified an association of smoking with future participation in shift work, but not of other cardiometabolic traits. This contrasts with our results, where we did not identify strong evidence for an effect of smoking heaviness on shift work selection. Given the inverse effect of educational attainment on shift work, and the known effect of educational attainment on reduced smoking^30^, it is possible that educational attainment confounded the previously observed relationship of smoking with shift work. Although shift workers typically report lower educational attainment^6,48^, prior work has not evaluated the role of educational attainment in shift work selection. Our findings suggest that this imbalance is driven by a causal effect of lower educational attainment on selection into shift work.

We observed a robust effect of higher BMI on shift work selection. This is in contrast to null associations in prior observational work^10,11^, There are several potential explanations for these differences. First, the observational estimates^10,11^ had wide confidence intervals, and could not exclude moderate effects of BMI on selection. Second, the influence of BMI on selection into shift work may accrue over time, such that effects are only observed later in life. Such a phenomenon would affect the present analysis, given the use of an older sample in contrast to the younger samples studied in prior studies of selection into shift work. However, this finding is consistent with the growing evidence for a causal effect of higher BMI on lower socioeconomic status^12,17^.

There are several potential pathways by which education and BMI may influence selection into shift work. Lower educational attainment is related to lower-skilled employment^49^, which is generally more typical of shift work^50^. It is less clear what mediates the effect of BMI on selection into shift work. One potential mechanism is weight stigma, which is a highly prevalent^51^, well-characterized driver of employment inequities^52^. Women experience a greater burden of this discrimination than men^53^, but we observed no interaction by sex, suggesting that men and women experience similar selection pressures into shift work on the basis of differences in adiposity and educational attainment. Although we hypothesized that differences in sleep timing preference due to variation in BMI^47^ or education may be a mediating mechanism, we did not observe an effect of earlier sleep timing preference on night shift odds. This is surprising given that night shift workers who tend to sleep earlier may experience the greatest burden of circadian misalignment^54,55^, and would therefore be more likely to select out of night shift work. A possible interpretation of this result is that selection effects of other social factors, such as socioeconomic status or educational attainment, preclude leaving a job on the basis of tolerability. We did not identify factors influencing selection out of shift work, but this sample was an order of magnitude smaller than the main sample, and we may have been underpowered to observe modest effects.

The present results have several potential implications for shift work research. The associations of shift work with cardiometabolic outcomes may be influenced by confounding by BMI and educational attainment. This does not invalidate the contribution of circadian misalignment to the excess disease risk conferred by shift work, but confounding may distort these estimates. Cross-sectional studies of shift work using obesity as an outcome may be at high risk for bias, as any observed associations will reflect a sum of forward and reverse causal effects. ^56^ Analyses restricted to shift workers may also be affected by collider bias^57^, which can induce false positive associations. Alternatively, shift work may mediate the effects of education and BMI on health outcomes. Recent MR mediation analyses showed that only half the variance of the effect of education on coronary artery disease risk is explained by conventional risk factors^30^, and shift work is an independent risk factor for cardiovascular disease^3^. Exposure to shift work may therefore explain part of the variance in this relationship. More broadly, MR may be a useful tool in occupational epidemiology to determine whether reverse causality may drive epidemiologic associations.

Our study has some limitations. We studied shift work in a middle-older aged population, and our results may therefore not generalize to younger populations. Given that UKB is a relatively healthy population^58^, our results may not generalize to sicker populations and may not be applicable to other populations or cohorts^59^ with different occupation structures or ethnicities. Moreover, the identified effects may be driven by selection bias in UKB given a 5% responder rate in the cohort^58^. Factors influencing selection into UKB may induce collider bias^57^ and false positive associations of exposures with selection into shift work. While we did not examine mediation through job-related factors, such as tolerability or fatigue, we were able to examine sleep timing preference and it did not appear to mediate the observed effects. Finally, despite consistent results in sensitivity analyses, the MR effect estimates may be biased by horizontal pleiotropy due to effects of genetic instruments on pathways influencing the outcome independently of the studied exposures. Triangulating results with analyses leveraging other forms of natural experiments, such as educational reform in the UK^60^, will strengthen confidence in causality.

In conclusion, higher BMI and lower educational attainment influenced selection into shift work and night shift work, while no other cardiometabolic risk factors were associated with selection into or out of shift work.

## Data Availability

The following data availability statement is made by the UK Biobank (https://www.ukbiobank.ac.uk/scientists-3/): “UK Biobank is an open access resource. The Resource is open to bona fide scientists, undertaking health-related research that is in the public good. Approved scientists from the UK and overseas and from academia, government, charity and commercial companies can use the Resource.”
Summary statistics used to generate the genetic instruments are all publicly available.

## Acknowledgements

This research has been conducted using the UK Biobank Resource (UK Biobank application number 6818). We would like to thank the participants and researchers from the UK Biobank who contributed or collected data.

